# Integrating CNS-active therapy with stereotactic radiotherapy for brain metastases: comparative outcomes of PULSAR and FSRT

**DOI:** 10.1101/2025.09.09.25335024

**Authors:** Michael Dohopolski, Lucian Zhao, Luiza Giuliani Schmitt, Lucas Mitre, Strahinja Stojadinovic, Michael Youssef, Evan Noch, Elizabeth Maher, Toral Patel, Ankur Patel, Matthew Sun, Hao Gao, Jiaxin Li, MinJae Lee, Robert Timmerman, Jill de Vis, Xin Cai, Tu Dan, Zabi Wardak

## Abstract

**Introduction:** Brain metastases (BMs) affect an increasing number of cancer patients and are typically managed with stereotactic radiosurgery (SRS). Our institution advocates the use of Personalized Ultrafractionated Stereotactic Adaptive Radiotherapy (PULSAR), where radiation is delivered in high-dose pulses at extended intervals allowing for treatment adaptation and easy concurrent systemic therapy integration. We explore the integration of PULSAR with central nervous system (CNS)-active drugs (CNS-aDs) compared to traditional fractionated stereotactic radiotherapy (FSRT).

**Methods:** This study involved a retrospective evaluation of patients treated with either PULSAR or FSRT using Gamma Knife from 2018-2024. We collected demographic, clinical, and specific treatment details, outcomes such as local failure (LF) and toxicity rates. Cumulative incidence analysis for local failure and toxicity, considering death a competing risk, and Kaplan-Meier survival analysis for overall survival (OS) were conducted. Multivariate (MV) Cox proportional hazard models assessed failure and toxicity rates.

**Results:** Analysis included 166 lesions treated with FSRT and 109 with PULSAR, predominantly in patients with lung and breast cancer. The median follow-up and OS were 1.88 and 1.48 years. 1- and 2-year LF rates were similar; 5%/8.9% vs 8.2%/12.3% for PULSAR and FSRT and 3.4%/5.5% vs 10.1%/12.3% with concurrent CNS-aDs (cCNS-aDs). BMs > 2 cm LF rates were 9.4% and 17.3% at two years for PULSAR and FSRT (p=0.1). No LFs were observed in BMs > 2 cm treated with PULSAR+CNS-aDs at 2.5 years. The two-year grade 3+ toxicity rate for PULSAR (8.7%) and FSRT (11.7%), without an increase in toxicity when combined with cCNS-aDs. BMs treated with PULSAR+cCNS-aDs were less likely to fail (HR 0.15, p=0.048) on MV analysis (MVA).

**Conclusion:** The integration of PULSAR with cCNS-aDs appears to offer excellent local control for larger brain metastases without increased toxicity. These promising results merit further prospective investigation to validate the findings and potentially establish new treatment protocols.

## 1. Introduction

Brain metastases are a common and challenging complication of systemic malignancies. Advances in radiation therapy, including stereotactic radiosurgery (SRS) and fractionated stereotactic radiotherapy (fSRT), have significantly improved the precision and efficacy of local treatment for brain metastases while minimizing damage to surrounding healthy brain tissue.

FSRT administers radiation over several sessions, typically spaced 1-3 days apart, improving local control of larger brain metastases to 80-90% while reducing adverse events^1,2^. However, FSRT’s effectiveness is limited by its static treatment plans, which do not account for tumor size changes during the treatment course. Some institutions have extended the interval between FSRT sessions to 2-4 weeks (historically called “Staged SRS”), allowing for treatment plan adjustments based on tumor response, which can reduce radiation exposure to healthy brain tissue^3–6^.

Our institution advocates Personalized Ultrafractionated Stereotactic Adaptive Radiotherapy (PULSAR), which uses ‘ultrafractionation’ to deliver high-dose radiation treatments PULSAR allows delivery of systemic therapy between PULSAR treatments, or pulses, which are oftentimes spaced 3-4 weeks apart. Immune checkpoint inhibitors (ICIs) and molecular targeted therapies have demonstrated substantial activity against various primary cancers and intracranial metastases^7–11^. Higher doses of radiation may synergize with ICI by amplifying the immune-mediated tumor response^12,13^.

Retrospective data have demonstrated that SRS in combination with immunotherapy have improved outcomes. For instance, in NSCLC, peri-operative administration of ICI in combination with SRS has demonstrated improved intracranial disease control compared to SRS alone^14^. In HER2-positive breast cancer, the concurrent use of anti-HER2 therapy or EGFR TKI with gamma knife SRS significantly reduced local failure rates^15,16^.

Unlike traditional SRS and FSRT combined with immunotherapy, which often increase adverse events, PULSAR may reduce them^17,18^. Preliminary studies suggest that PULSAR may offer synergistic benefits, potentially transforming BM management into a more personalized and effective regimen with improved tolerability^13^. The present study investigates whether PULSAR or fSRT provides superior local control and comparable toxicity when administered with a concurrent CNS-active drug (cCNS-aD).

## 2. Methods

## 2A. Patient Selection

Patients with BMs treated with PULSAR or FSRT using Gamma Knife between January 2018 and March 2024 were retrospectively evaluated^19^. A total of 412 lesions across 322 patients were identified. 235 lesions in 147 patients treated from February 2018 to January 2024 received FSRT; 177 lesions in 116 patients received PULSAR starting from May 2020 to June 2024. Subsequent chart reviews verified treatment intent, excluding post-operative radiation therapy (PORT), and ensured a minimum of four-month follow-up post-treatment (but still including patients deceased within this timeframe). This process resulted in the inclusion of 166 lesions (92 patients) in the FSRT cohort. Exclusions from the FSRT group comprised 19 lesions with inadequate follow-up, 15 non-BM lesions, 34 lesions treated with PORT, and one lesion receiving a single-fraction treatment. For the PULSAR cohort, 109 lesions (55 patients) were eligible. Exclusions encompassed 24 lesions with insufficient follow-up, 25 non-BM lesions, 15 lesions initially planned for PULSAR but ultimately not treated, and 4 lesions receiving PORT. (**Supplemental Figure B1**).

## 2B. PULSAR Treatments

Our group previously defined PULSAR as using large dose “pulses” separated by weeks or months, allowing for observation and adaptive replanning based on significant changes in the tumor and its environment for more personalized and effective therapy^13^. In this study, PULSAR regimens varied with 2-4 week intervals between treatments, necessitating repeat magnetic resonance imaging (MRI) to assess the need for replanning. After each MRI, the treating physician determined whether to adapt or continue the treatment plan. The choice of regimen is physician-dependent, influenced by factors such as tumor size, prior radiotherapy, histology, and critical organ involvement. We observed that many patients experienced the most significant decrease in tumor response within the first month. Consequently, we often use the Triple Threat approach to deliver most of the radiation dose before the break, maximizing initial tumor impact and allowing for clinically significant treatment adaptations. Patients received either two or five treatments.

**PULSAR Styles:**

- **2 Adaptations, q4wks:** Two adaptive replanning sessions with a four-week interval between each treatment. The specific regimen involves 15 Gy, a 4-week break, and then another 15 Gy.
- **Triple Threat:** Three treatments are delivered every other day, followed by a 3-4 week break, then two final treatments every other day. This is the only technique that starts with a traditional fractionated approach, then pauses for a rescan before finishing within 3-4 weeks. For example, 5-6 Gy x 3 every other day, then a 3-4 week break, followed by 5-7.5 Gy x 2 every other day.
- **4 Adaptations, varying intervals:** Four adaptations with varying intervals. For example, 6 Gy x 5, adapted for 4 of the 5 treatments with variably spaced treatments.
- **4 Adaptations, q4wks:** Four adaptations with fixed four-week intervals. For example, 6 Gy x 5, adapted for 4 of the 5 treatments, spaced 4 weeks apart.
- **5 Adaptations, q2wks:** Five adaptations with two-week intervals. For example, 6 Gy x 5, adapted for all 5 treatments, spaced 2 weeks apart.
- **5 Adaptations, q3wks:** Five adaptations with three-week intervals. For example, 6 Gy x 5, adapted for all 5 treatments, spaced 3 weeks apart.
- **5 Adaptations, q4wks:** Five adaptations with four-week intervals. For example, 6 Gy x 5, adapted for all 5 treatments, spaced 4 weeks apart.

To minimize toxicity, nearest neighbor targets treated with single SRS prescriptions are not treated on the same day and are optimally distributed throughout the treatment course^20^.

### 2C. Clinical Features

Patient characteristics recorded included age at diagnosis, histology, history of prior radiation therapy (specified per lesion), dates of clinical and MRI follow-ups, and death dates where applicable. Treatment details included concurrent (administered during radiation treatment or within a week following its conclusion) or adjuvant systemic therapies, radiation treatment dates, doses, and fraction numbers. Recurrence patterns (in-field or marginal) and treatment-related toxicities were systematically documented. Diagnoses were made in a multidisciplinary setting after reviewing serial imaging, with at least two physicians from neuroradiology, neuro-radiation oncology, neurosurgery, and neuro-oncology.

Local failures were classified as in-field or marginal, with in-field failures occurring within the prescription volume and marginal failures identified as asymmetric recurrences at a single edge of the prescription volume.

A list of CNS-aDs is provided in **Supplemental Table A1**.

### 2D. Analyses

Clinical and treatment-related variables among patients undergoing FSRT versus PULSAR were compared using the *χ*^2^ test or Fisher exact test for categorical variables and the 2-sample t-test or Mann-Whitney U test for continuous variables, as appropriate.

To evaluate local failure and grade 3+ vasogenic edema, we performed competing risk regression (CRR) and calculated cumulative incidence, with death as a competing risk using *tidycmprsk* (R package). Gray’s test was used to assess the statistical significance. Multivariate Cox proportional hazard models were constructed for time to local failure and time to grade 3+ vasogenic edema, incorporating clinically relevant variables, using *survival* (R package). OS was analyzed using the Kaplan-Meier method, with the log-rank test employed to compare survival distributions among different groups.

To account for the clustered nature of the data, where patients may have multiple brain metastases treated, we performed repeated analyses for CRR and Cox proportional hazard models using the *crrSC* and *frailtypack* R packages, respectively.

Variables included in LF MVA: Treatment Type (FSRT/PULSAR), aCNS-aD (no/yes), cCNS-aD (no/yes), BED (continuous, Gy), Radioresponse (Radiosensitive/Radioresistant), Initial Size (continuous, cm^3^, Age (continuous, years).

Variables included in grade 3+ vasogenic edema MVA: Treatment Type (FSRT/PULSAR), aCNS-aD (no/yes), cCNS-aD (no/yes), BED (continuous, Gy), Prior RT (no/yes), Radioresponse (Radiosensitive/Radioresistant), Initial Size (continuous, cm^3^, Age (continuous, years).

We included an interaction term (Treatment Type:cCNS-aD) in both analyses to explore whether the effect of Treatment Type (FSRT/PULSAR) on the hazard rate is different depending on whether cCNS-aD is present or not.

All statistical analyses were conducted using *R (version 4.3.3)*.

## 3. Results

### 3A. Patient Demographics and Treatment Parameters

The median age for patients receiving FSRT was 64 years old (54-72 years old). Non-small cell lung cancer (NSCLC) (34%), breast cancer (19%), and gastrointestinal (GI) (9.0%) were the most common histologies (**Table 1**). A significant proportion (71%) did not receive cCNS-aDs, and 70% did receive adjuvant CNS-aD (aCNS-aD) therapy. Conversely, patients receiving PULSAR had a slightly lower median age of 61 years old (56-67 years old, p=0.4). NSCLC (37%) and breast cancer (28%) were the most prevalent histologies. cCNS-aD use was higher (61%, p<0.0001). Prior radiation therapy and radioresponsive histologies were similar between groups. More BMs in the PULSAR cohort were treated with 6 Gy or above (61.6% vs. 88.6%, p < 0.001). Fifty-one (51) lesions were greater than 2 cm in diameter.

**Table 1.** Patient and lesion demographic and treatment characteristics. Age was calculated per person, while other variables were calculated per lesion. *Abbreviations: PULSAR, Personalized Ultrafractionated Stereotactic Adaptive Radiotherapy; FSRT, Fractionated Stereotactic Radiotherapy; GU, genitourinary; GI, gastrointestinal; Gyn, gynecological; NSCLC, non-small cell lung cancer; cCNS-aD, concurrent central nervous system-active drug; aCNS-aD, adjuvant central nervous system-active drug; RT, radiation therapy; SRS, stereotactic radiosurgery; Gy, gray; cGy, centiGy; fx, fraction; q2wks, every two weeks; q3wks, every three weeks; q4wks, every four weeks.* *Adaptations include initial plan plus addition adaptive plans.

| Variables | FSRT, N (lesions) = 166 | PULSAR, N (lesions) = 109 | p-value |
| --- | --- | --- | --- |
| **Age (yr)** |  |  |  |
|  | 64 (54, 72) | 62 (58, 68) | 0.4 |
| **Initial Size (cm<sup>3</sup>)** |  |  |  |
|  | 3 (1, 9) | 4 (1, 9) | 0.6 |
| **Histology** |  |  |  |
| GU | 14 (8.4%) | 13 (12%) | — |
| Breast | 32 (19%) | 30 (28%) |  |
| GI | 15 (9.0%) | 5 (4.6%) |  |
| Gyn | 14 (8.4%) | 4 (3.7%) |  |
| Melanoma | 10 (6.0%) | 12 (11%) |  |
| NSCLC | 57 (34%) | 40 (37%) |  |
| Small cell | 10 (6.0%) | 1 (0.9%) |  |
| Other | 14 (8.4%) | 4 (3.7%) |  |
| **Radioresponse** |  |  |  |
| Radioresistant | 46 (28%) | 31 (28%) | 0.9 |
| Radiosensitive | 120 (72%) | 78 (72%) |  |
| **PULSAR Style\*** |  |  |  |
| 2 Adaptations, q4 weeks | - | 1 (0.9%) | Not Applicable |
| Triple Threat | - | 94 (86%) |  |
| 4 Adaptations, varying intervals | - | 3 (2.8%) |  |
| 4 Adaptations, q4 weeks | - | 1 (0.9%) |  |
| 5 Adaptations, q2 weeks | - | 3 (2.8%) |  |
| 5 Adaptations, q3 weeks | - | 5 (4.6%) |  |
| 5 Adaptations, q4 weeks | - | 1 (0.9%) |  |
| **cCNS-aD** |  |  |  |
| No | 116 (70%) | 43 (39%) | <0.001 |
| Yes | 50 (30%) | 66 (61%) |  |
| **aCNS-aD** |  |  |  |
| No | 52 (31%) | 20 (18%) | 0.017 |
| Yes | 114 (69%) | 89 (82%) |  |
| **cCNS-aD and aCNS-aD** |  |  |  |
| No | 116 (70%) | 47 (43%) | <0.001 |
| Yes | 50 (30%) | 62 (57%) |  |
| **Any Concurrent Systemic Therapy** |  |  |  |
| No | 116 (70%) | 36 (33%) | <0.001 |
| Yes | 50 (30%) | 73 (67%) |  |
| **Any Adjuvant Systemic Therapy** |  |  |  |
| No | 44 (27%) | 16 (15%) | 0.020 |
| Yes | 122 (73%) | 93 (85%) |  |
| **Concurrent and Adjuvant Systemic Therapy** |  |  |  |
| No | 116 (70%) | 39 (36%) | <0.001 |
| Yes | 50 (30%) | 70 (64%) |  |
| **Prior RT** |  |  |  |
| No | 134 (81%) | 94 (86%) | 0.2 |
| Yes | 32 (19%) | 15 (14%) |  |
| **Total Dose (cGy)** |  |  |  |
| Dose | 3,000 (2,500, 3,000) | 3,000 (3,000, 3,000) | 0.016 |
| **Number of Fractions** |  |  |  |
| Fractions | 5.00 (5.00, 5.00) | 5.00 (5.00, 5.00) | 0.012 |
| **Dose per Fraction (cGy/fx)** |  |  |  |
| 400 | 7 (4.2%) | 0 (0%) | <0.001 |
| 500 | 45 (27%) | 10 (9.2%) |  |
| 550 | 11 (6.6%) | 2 (1.8%) |  |
| 600–700 | 103 (61.6%) | 95 (86.8%) |  |
| 1500 | 0 (0%) | 2 (1.8%) |  |
| **BED (Gy)** |  |  |  |
|  | 48.0 (37.5, 48.0) | 48.0 (48.0, 48.0) | 0.002 |

### 3B. OS

The median follow-up (FU) duration was 1.88 years (interquartile range (IQR), 1.30-2.74 years). The median OS for the entire cohort was 1.48 years (IQR, 0.56-3.66 years), as depicted in **Supplemental Figure B2A**. Stratification of OS by FSRT vs. PULSAR, other vs. PULSAR+cCNS-aD use, or PULSAR alone (both with and without a cCNS-aD use) revealed no significant differences. Refer to **Supplemental Figure B2B-D** for detailed comparisons.

### 3C. Cumulative Incidence and Case-Specific Multivariate Cox Proportional Hazard Model - Local Failure

The FSRT cohort experienced 20 local failures (18 patients), with cumulative incidence rates increasing from 8.2% at one year to 12.3% by the second year. Seven local failures (7 patients) were noted in the PULSAR cohort, with incidence rates growing from 5.0% at one year to 8.9% by the second year. No significant difference in local failure rates was observed between these groups (Gray’s test, p=0.21, **Figure 1A**). BMs treated with PULSAR+cCNS-aDs exhibited lower failure rates–3.4% and 5.5% at one and two years compared to those treated with FSRT+cCNS-aDs, which had rates of 10.1% and 12.3%; however, these differences were not statically different. **Figure 1C**). The difference in local failure rates was more pronounced for lesions larger than 2cm—9.4% and 17.3% at two years for PULSAR and FSRT, respectively (**Figure 1B**, p=0.10). Notably, there were no local failures among BMs larger than 2cm treated with PULSAR+cCNS-aDs at 2.5 years (**Figure 1D**).

**Figure 1:**
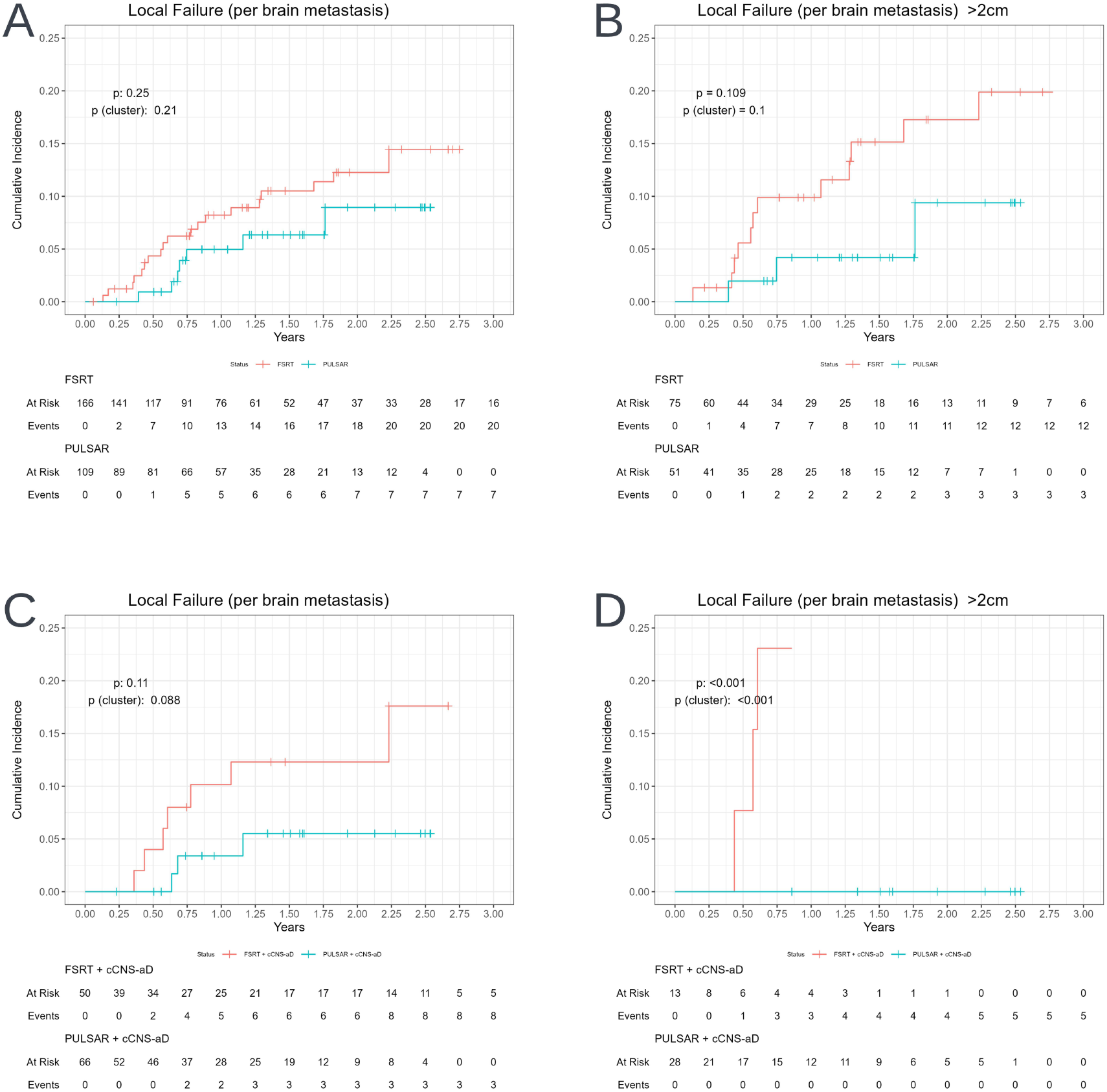
**A)** Cumulative incidence of local failure for the entire cohort. **B)** Cumulative incidence of local failures for lesions > 2 cm in diameter. **C)** Cumulative incidence of local failure for the entire cohort stratified by cCNS-aD. **D)** Cumulative incidence of local failures for lesions > 2 cm in diameter stratified by cCNS-aD. *Abbreviations: FSRT, Fractionated Stereotactic Radiotherapy; PULSAR, Personalized Ultrafractionated Stereotactic Adaptive Radiotherapy; cCNS-aD, Concurrent Central Nervous System-Active Drug*.

The case-specific multivariate Cox proportional hazard model identified that aCNS-aD use and radiosensitive were associated with reduced local failure rates (p<0.05). Conversely, cCNS-aD use was linked to an increased risk of local failure. A trend towards an increased risk of failure with larger lesion size was observed. Trending toward significance, the interaction between PULSAR and cCNS-aD was associated with reduced local failure rates (**Table 2A**, p=0.053). Overall, the clustered data-based analyses supported these observations, but the interaction between PULSAR and cCNS-aD showed a significant reduction in local failures **Table 2B**, p=0.048). Local failures were more frequent with cCNS-aD use in the FSRT group. Details of the FSRT cohort receiving cCNS-aDs are provided in **Supplemental Table A2**.

**Table 2.** Case-specific multivariate Cox proportional hazard model for local failure, incorporating treatment characteristics, tumor characteristics, and patient factors. **A)** Non-clustered data analysis. **B)** Clustered data analysis (ie patients may have multiple brain metastases treated). *Abbreviations: PULSAR, Personalized Ultrafractionated Stereotactic Adaptive Radiotherapy; cCNS-aD, concurrent central nervous system-active drug; aCNS-aD, adjuvant central nervous system-active drug; HR, hazard ratio; CI, confidence interval*.

| Variables | A |  |  | B |  |  |
| --- | --- | --- | --- | --- | --- | --- |
|  | HR | 95% CI | p-value | HR | 95% CI | p-value |
| **Treatment Characteristics** |  |  |  |  |  |  |
| *Treatment Type (PULSAR)* | 1.26 | [0.40, 4.00] | 0.695 | 1.26 | [0.41, 3.86] | 0.686 |
| *aCNS-aD (aCNS-aD)* | 0.24 | [0.09, 0.63] | 0.004 | 0.24 | [0.09, 0.64] | 0.005 |
| *cCNS-aD (cCNS-aD)* | 5.89 | [1.62, 21.37] | 0.007 | 5.89 | [1.65, 21.02] | 0.006 |
| *BED* | 1.03 | [0.97, 1.11] | 0.347 | 1.03 | [0.97, 1.10] | 0.329 |
| *Treatment Type(PULSAR)\*cCNS-aD (cCNS-aD)* | 0.13 | [0.02, 1.02] | 0.053 | 0.13 | [0.02, 0.98] | 0.048 |
| **Tumor Characteristics** |  |  |  |  |  |  |
| *Radioresponse* |  |  |  |  |  |  |
| Radiosensitive | 0.43 | [0.19, 0.94] | 0.035 | 0.43 | [0.19, 0.96] | 0.039 |
| *Initial Size* | 1.03 | [0.98, 1.08] | 0.175 | 1.03 | [0.98, 1.08] | 0.192 |
| **Patient Factors** |  |  |  |  |  |  |
| *Age (yr)* | 1.03 | [0.99, 1.06] | 0.117 | 1.03 | [0.99, 1.06] | 0.117 |

### 3D. Cumulative Incidence and Case-Specific Multivariate Cox Proportional Hazard Model - Toxicity

The FSRT cohort experienced 19 grade 3+ toxicity events (15 patients), while the PULSAR group reported 6 grade 3+ toxicity events (6 patients). The cumulative incidence of toxicity in the FSRT group rose from 8.1% in one year to 11.7% by the second year. In contrast, the PULSAR group’s toxicity rate remained steady at 8.7% over two years. No significant difference was observed in toxicity rates between the two groups (p=0.49, **Figure 2A**). **Figure 2C** shows the cumulative incidence of grade 3+ vasogenic edema stratified by cCNS-aD use for both FSRT and PULSAR groups. The two-year rates were 4.8% for PULSAR+cCNS-aD use and 6.4% for FSRT+cCNS-aD use cohorts. The two-year rates for lesions > 2cm were 14.6% and 14.5% for the treatment group (FSRT vs. PULSAR, **Figure 2B**) and 7.7% vs. 3.6% for treatment group + cCNS-aDs (**Figure 2D**).

**Figure 2:**
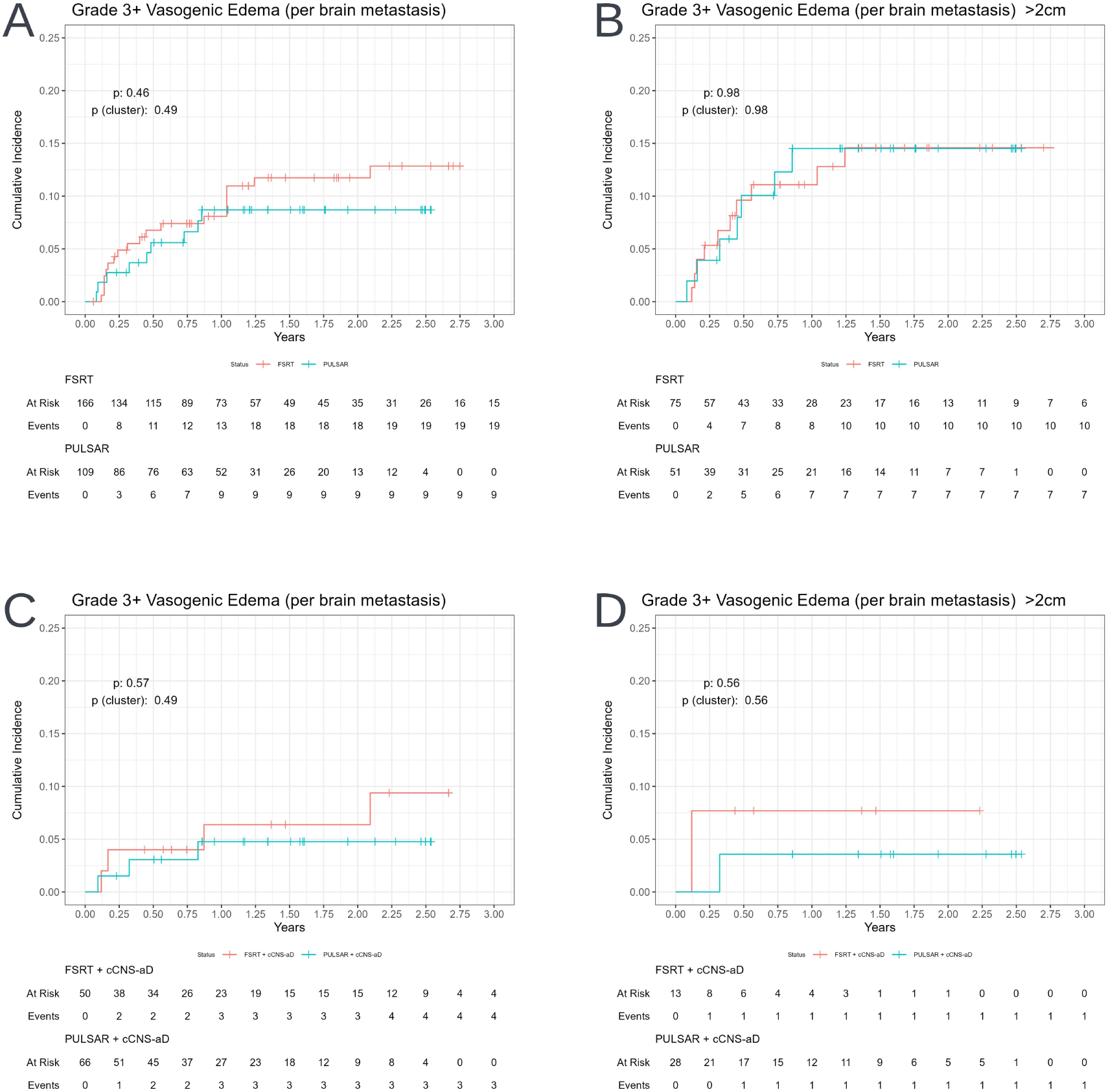
**A)** Cumulative incidence of grade 3+ vasogenic edema for the entire cohort. **B)** Cumulative incidence of grade 3+ vasogenic edema for lesions > 2 cm in diameter. **C)** Cumulative incidence of grade 3+ vasogenic edema for the entire cohort stratified by cCNS-aD.**D)** Cumulative incidence of grade 3+ vasogenic edema for lesions > 2 cm in diameter stratified by cCNS-aD. *Abbreviations: FSRT, Fractionated Stereotactic Radiotherapy; PULSAR, Personalized Ultrafractionated Stereotactic Adaptive Radiotherapy; cCNS-aD, Concurrent Central Nervous System-Active Drug*.

The case-specific multivariate Cox proportional hazard model without and with clustering did not have any variables associated with vasogenic edema (**Table 3**).

**Table 3.** Case-specific multivariate Cox proportional hazard model assessing grade 3+ vasogenic edema, incorporating treatment characteristics, tumor characteristics, and patient factors. **A)** Non-clustered data analysis. **B)** Clustered data analysis (ie patients may have multiple brain metastases treated). *Abbreviations: PULSAR, Personalized Ultrafractionated Stereotactic Adaptive Radiotherapy; aCNS-aD, adjuvant central nervous system-active drug; cCNS-aD, concurrent central nervous system-active drug; RT, radiation therapy; HR, hazard ratio; CI, confidence interval*.

| Variables | A |  |  | B |  |  |
| --- | --- | --- | --- | --- | --- | --- |
|  | HR | 95% CI | p-value | HR | 95% CI | p-value |
| **Treatment Characteristics** |  |  |  |  |  |  |
| *Treatment Type (PULSAR)* | 1.14 | [0.42, 3.09] | 0.794 | 1.14 | [0.34, 3.86] | 0.831 |
| *aCNS-aD (aCNS-aD)* | 1.47 | [0.58, 3.73] | 0.419 | 1.47 | [0.54, 3.99] | 0.451 |
| *cCNS-aD (cCNS-aD)* | 0.76 | [0.19, 2.99] | 0.690 | 0.76 | [0.17, 3.41] | 0.716 |
| *BED* | 1.07 | [0.96, 1.20] | 0.211 | 1.07 | [0.96, 1.19] | 0.203 |
| *Prior RT* | 1.31 | [0.36, 4.73] | 0.678 | 1.31 | [0.37, 4.62] | 0.672 |
| *Treatment Type(PULSAR)\*cCNS-aD (cCNS-aD)* | 0.33 | [0.05, 2.02] | 0.232 | 0.33 | [0.05, 2.40] | 0.275 |
| **Tumor Characteristics** |  |  |  |  |  |  |
| *Radioresponse* |  |  |  |  |  |  |
| Radiosensitive | 0.62 | [0.26, 1.48] | 0.282 | 0.62 | [0.20, 1.90] | 0.402 |
| *Initial Size (cm<sup>3</sup>)* | 1.03 | [0.99, 1.08] | 0.178 | 1.03 | [0.98, 1.08] | 0.228 |
| **Patient Factors** |  |  |  |  |  |  |
| *Age (yr)* | 1.01 | [0.98, 1.04] | 0.557 | 1.01 | [0.97, 1.05] | 0.625 |

**Table 4.** Relative change in volume at the final adaptive “Pulse” for the PULSAR cohort, stratified by initial tumor size, cCNS-aD use, PULSAR style, and histology. Median, 25th, and 75th percentile values are presented for each category. *Abbreviations: PULSAR, Personalized Ultrafractionated Stereotactic Adaptive Radiotherapy; cCNS-aD, concurrent central nervous system-active drug; q2wks, every two weeks; q3wks, every three weeks; q4wks, every four weeks; NSCLC, non-small cell lung cancer; GI, gastrointestinal; GU, genitourinary. GYN, gynecological.* *Adaptations include initial plan plus additional adaptive plans.

| Category | Median (%) | 25 (%) | 75 (%) |
| --- | --- | --- | --- |
| **General Volume Reduction** |  |  |  |
| Small (<2 cm) | 34.2 | 0.0 | 59.6 |
| Medium (2-4 cm) | 38.1 | 7.7 | 57.3 |
| Large (>4 cm) | 31.3 | 23.3 | 35.3 |
| **Volume Reduction by cCNS-aD use** |  |  |  |
| *Small (<2 cm)* |  |  |  |
| No | 0.0 | 0.0 | 31.7 |
| Yes | 49.0 | 32.4 | 64.2 |
| *Medium (2-4 cm)* |  |  |  |
| No | 28.9 | 0.0 | 51.8 |
| Yes | 49.2 | 27.6 | 65.1 |
| *Large (>4 cm)* |  |  |  |
| No | 23.3 | 19.3 | 27.3 |
| Yes | 39.3 | 39.3 | 39.3 |
| **Volume Reduction by PULSAR Style\*** |  |  |  |
| *Small (<2 cm)* |  |  |  |
| Triple Threat | 32.4 | 0.0 | 54.6 |
| 2 Adaptations, q4wks | 3.3 | 3.3 | 3.3 |
| 4 Adaptations, varying intervals | 79.9 | 78.7 | 81.1 |
| 5 Adaptations, q4wks | -50.0 | -50.0 | -50.0 |
| 5 Adaptations, q3wks | 37.0 | 18.5 | 39.1 |
| 5 Adaptations, q2wks | 63.4 | 60.8 | 66.1 |
| *Medium (2-4 cm)* |  |  |  |
| Triple Threat | 38.1 | 7.7 | 57.3 |
| 4 Adaptations, varying intervals | 85.4 | 85.4 | 85.4 |
| 5 Adaptations, q3wks | 30.8 | 29.2 | 32.4 |
| 4 Adaptations, q4wks | -2.8 | -2.8 | -2.8 |
| *Large (>4 cm)* |  |  |  |
| Triple Threat | 27.3 | 21.3 | 33.3 |
| 5 Adaptations, q2wks | 31.3 | 31.3 | 31.3 |
| **Volume Reduction by Histology** |  |  |  |
| *Small (<2 cm)* |  |  |  |
| NSCLC | 0.0 | 0.0 | 52.5 |
| Breast | 35.3 | 11.4 | 57.6 |
| GI | 44.4 | 44.4 | 44.4 |
| Melanoma | 50.3 | 50.3 | 50.3 |
| GU | 44.7 | 17.8 | 63.4 |
| GYN | 85.6 | 85.6 | 85.6 |
| Other | 61.2 | 61.2 | 61.2 |
| *Medium (2-4 cm)* |  |  |  |
| NSCLC | 51.7 | 5.6 | 64 |
| Breast | 31.5 | 19.2 | 37.2 |
| GI | 23.9 | 0.0 | 52.9 |
| Melanoma | 12.7 | -5.5 | 31.0 |
| GU | 38.1 | 38.1 | 38.1 |
| GYN | 52.2 | 40.5 | 76.1 |
| Other | 24.7 | 11.5 | 48.9 |
| *Large (>4 cm)* |  |  |  |
| NSCLC | 39.3 | 39.3 | 39.3 |
| Breast | 23.3 | 19.3 | 27.3 |

## 4. Discussion

The integration of stereotactic radiotherapy with CNS-aDs is now common in practice.^21,22^ In this context, we compared PULSAR with conventionally spaced FSRT in a modern cohort (2021–2024) in which targeted therapies, antibody-based agents, and immunotherapies were used. Across 109 BMs treated with PULSAR and 166 with FSRT (median follow-up ∼1.9 years), PULSAR used with concurrent or adjuvant CNS-aDs was associated with numerically low rates of LF without an observed increase in high-grade toxicity compared to FSRT. Multivariable analyses further suggested associations between CNS-aD use, radioresponsive histologies, and lower failure, with an interaction pattern between treatment type (PULSAR) and cCNS-aD consistent with synergy, although the study was not powered for definitive interaction testing. These findings are hypothesis-generating and suggest potential benefit warranting further study.

Our findings are consistent with the fractionation literature emphasizing dose and lesion size. In a systematic review, Lehrer etal. summarized multiple retrospective studies where FSRT achieved ∼90% local control with acceptable toxicity.^1^ Minniti etal. (2016) reported superior 1-year local control for FSRT versus single-fraction SRS in untreated BMs > 2cm (91% vs. 77%) and lower RN (8% vs. 20%).^2^ The international multicenter study by Ehret et al. in brainstem metastases showed excellent 12- and 24-month local control (82.9% and 71.4%), with no significant difference between SRS and FSRT after accounting for biologically effective dose (BED); higher BED correlated with improved control.^23^

Evidence supporting the effectiveness of pairing local therapy with CNS-active modern agents continues to grow, particularly in disease-specific contexts. In HER2^+^ breast cancer, the TROG16.02 *LOCAL HERO* phaseII study reported a 12-month freedom from local failure of 91% with low toxicity, and 95% avoidance of WBRT at one year when SRS and/or surgery were delivered with HER2-directed therapy.^15^ A complementary multi-institutional series of trastuzumab deruxtecan (T-DXd) with SRS demonstrated 12-month local control of 97% across 215 lesions, with a 24-month symptomatic radionecrosis incidence of 2.1% per lesion (11% per patient), while highlighting worse distant CNS control for HER2-low disease.^16^ Beyond HER2-directed regimens, contemporary reports suggest favorable interactions between stereotactic radiation and immune checkpoint inhibitors (ICIs) or CNS-penetrant TKIs, though the optimal timing remains unresolved.^21,22,24^ In NSCLC BMs, Dohm et al. observed improved local and distant intracranial control when systemic therapy was delivered concurrently or upfront with SRS versus delayed therapy, without a significant increase in radionecrosis (RN).^25^ Taken together, these data support integrating effective local therapy with active systemic regimens. Balanced against these efficacy signals, safety considerations with SRS/FSRT under active systemic therapy are nontrivial. With ICIs, higher PD-L1 expression has been associated with increased grade ≥2 RN when ICIs are delivered near SRS/SRT, and symptomatic RN has been reported in several series.^17,18^ For antibody–drug conjugates (ADCs), multicenter analyses and meta-analyses suggest clinically meaningful RN risk with concurrent therapy—particularly for intracranial targets—with T-DM1 frequently cited.^26–28^ Collectively, these reports suggest risk is drug- and timing-dependent and underscore the need to define safe integration windows.

Our PULSAR cohort—including patients receiving cCNS-aDs—did not show excess high-grade toxicity relative FSRT. Observed rates fell within published ranges for staged/ultrafractionated approaches (typically 4–11%, with some series up to ∼18%).^3,5,29–32^ Importantly, we did not observe the elevated RN signals reported in some ICI- or ADC-concurrent SRS/FSRT series. While our study was not powered for drug-class–specific analyses, the absence of excess toxicity despite frequent cCNS-aD use is consistent with the hypothesis that time-separated pulses and on-treatment replanning may limit harmful dose–drug overlaps; whether PULSAR’s spacing is protective, neutral, or harmful remains unknown and warrants prospective evaluation. Where PULSAR may be distinctive is in using schedule as a therapeutic lever. Multi-week pulses permit sustained on-treatment drug exposure and mid-course adaptation based on early volumetric response. If schedule-dependent synergy exists, it should manifest most clearly in larger metastases—where hypoxia, edema, and normal-tissue constraints narrow therapeutic margins—the subgroup in which our signal was most evident (≥ 2cm). By distributing dose in time and enabling adaptive replanning, PULSAR may preserve the benefits of integration while potentially reducing harmful spatiotemporal dose–drug overlaps that underlie RN in some settings.

Our findings extend earlier work on ultrafractionated or staged approaches, which achieved high local control in large BMs but largely predated widespread CNS-aD use or did not report it explicitly.^4–6,29–31,33^ Conversely, while modern series increasingly support concurrent systemic therapy with SRS/FSRT—particularly ICIs and CNS-penetrant TKIs—whether an ultrafractionated pulse strategy modifies benefit relative to closely spaced FSRT remains undetermined.^21,22,24,34^ By directly comparing PULSAR and FSRT in a CNS-aD era, our analysis targets this scheduling question rather than a simple modality comparison.

This study has limitations inherent to retrospective, single-institution analyses. Treatment selection, planning, and adaptation were clinician-dependent. The PULSAR cohort had greater CNS-aD use and, in some cases, higher biologically effective dose, both of which could confound local control. Adjuvant therapy was common across groups (∼70–80%), and time-dependent exposure introduces potential immortal-time bias. Toxicity adjudication was not blinded, and the sample was insufficient for drug class-specific analyses or precise interaction estimates.

## 5. Conclusion

In a contemporary cohort with frequent CNS-active systemic therapy, PULSAR achieved low local failure without an observed increase in high-grade toxicity relative to FSRT, with the clearest signal in larger lesions (≥ 2,cm). While therapy-specific radionecrosis has been reported when SRS/FSRT is delivered near certain agents, we did not observe comparable excess toxicity with PULSAR. Moreover, data suggest superior local control when concurrent CNS-active systemic therapy was paired with PULSAR, generating the hypothesis that temporal spacing and on-treatment adaptation may at least preserve and might even improve efficacy while not increasing toxicity. Prospective, drug-class–aware trials are needed to determine whether pulsed scheduling confers benefit beyond dose and drug selection and to define safe integration windows with modern CNS-active therapies.

## Data Availability

All data produced in the present study are available upon reasonable request to the authors.

## Supplement A

**Table A1.**
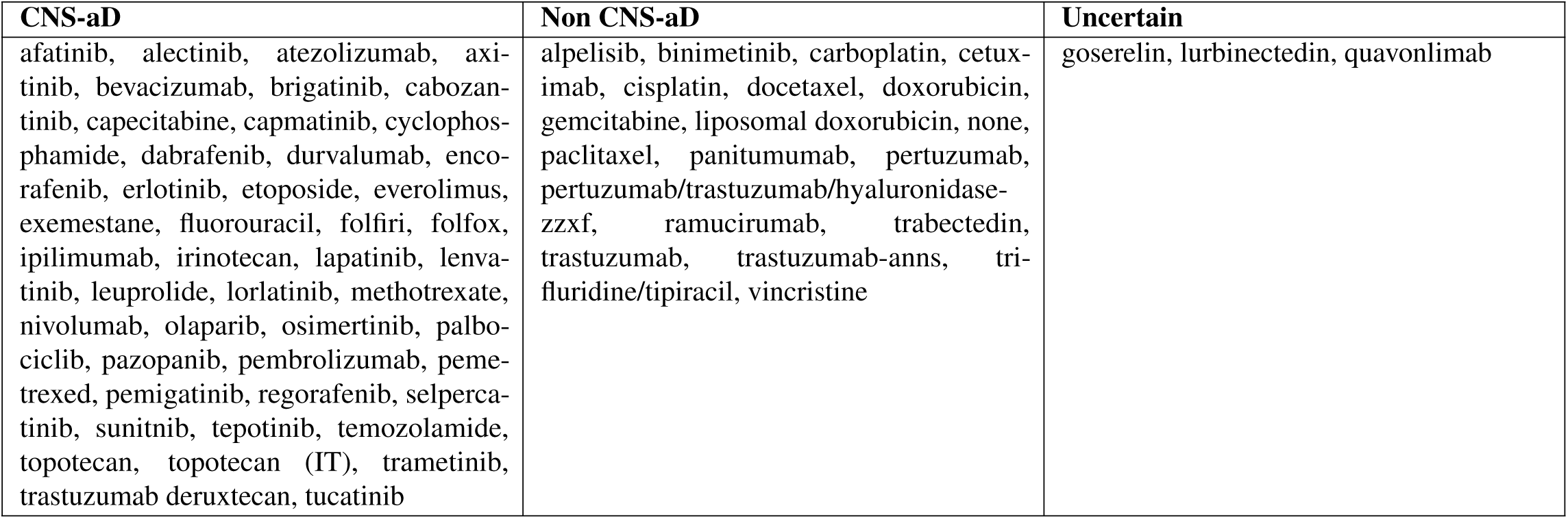
Central nervous system-active drug (CNS-aD). Therapeutic agents were included in the CNS-aD cohort if they have a reported history intracranial activity in the literature. Drugs in the uncertain cohort were included in the non CNS-aD cohort. Multidrug regimens were included in CNS-aD cohort if a single agent had reported intracranial activity. Intrathecal therapies were included in the CNS-aD cohort.

**Table A2.**
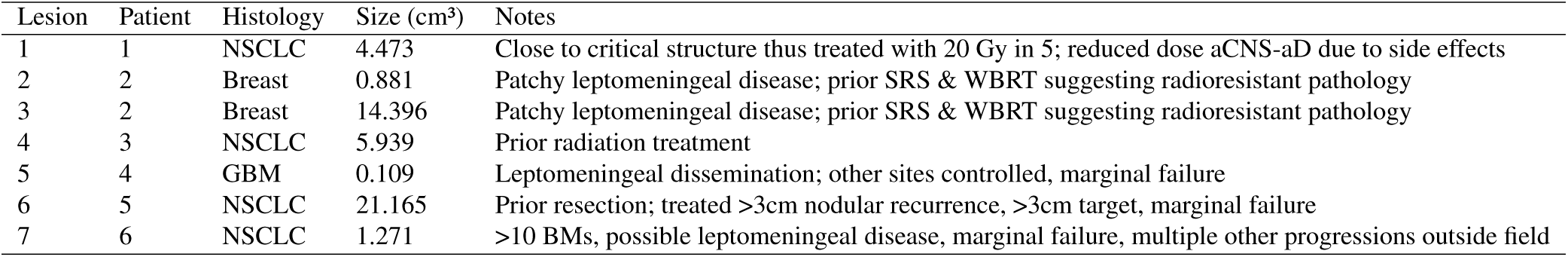
Summary of BMs treated with cCNS-aDs and FSRT. This exploration was prompted by an increased risk of local failure noted in the multivariate model, with the largest proportion of failures within this cohort. The table includes details on lesion characteristics, patient information, and relevant treatment notes. *Abbreviations: BMs, brain metastases; FSRT, fractionated stereotactic radiotherapy; NSCLC, non-small cell lung cancer; GBM, glioblastoma; aCNS-aD, adjuvant central nervous system-active drug; SRS, stereotactic radiosurgery; WBRT, whole brain radiotherapy*.

| Lesion | Patient | Histology | Size (cm<sup>3</sup>) | Notes |
| --- | --- | --- | --- | --- |
| 1 | 1 | NSCLC | 4.473 | Close to critical structure thus treated with 20 Gy in 5; reduced dose aCNS-aD due to side effects |
| 2 | 2 | Breast | 0.881 | Patchy leptomeningeal disease; prior SRS & WBRT suggesting radioresistant pathology |
| 3 | 2 | Breast | 14.396 | Patchy leptomeningeal disease; prior SRS & WBRT suggesting radioresistant pathology |
| 4 | 3 | NSCLC | 5.939 | Prior radiation treatment |
| 5 | 4 | GBM | 0.109 | Leptomeningeal dissemination; other sites controlled, marginal failure |
| 6 | 5 | NSCLC | 21.165 | Prior resection; treated >3cm nodular recurrence, >3cm target, marginal failure |
| 7 | 6 | NSCLC | 1.271 | >10 BMs, possible leptomeningeal disease, marginal failure, multiple other progressions outside field |

## Supplement B

**Figure B1.**
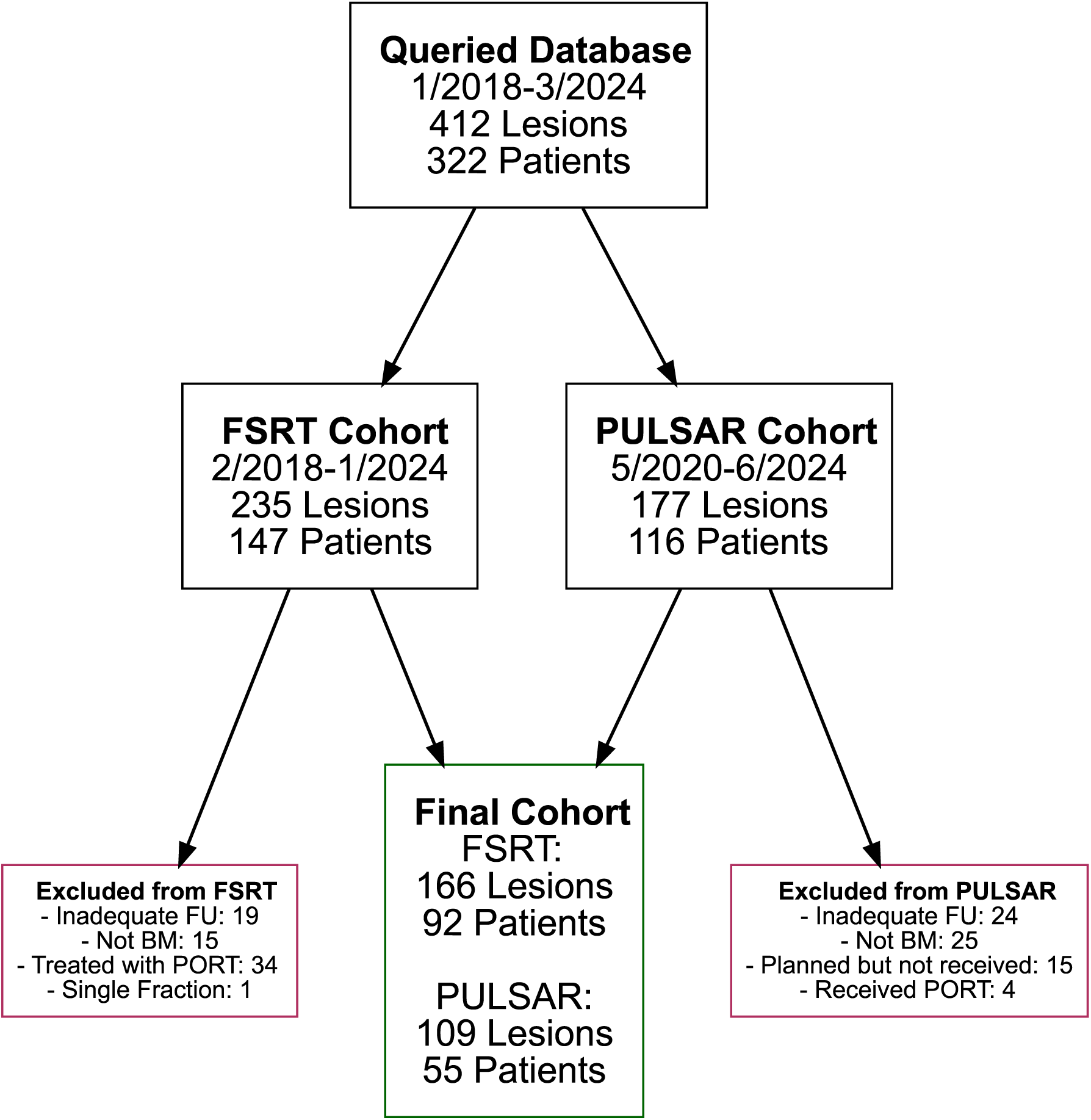
Consort diagram.

**Figure B2.**
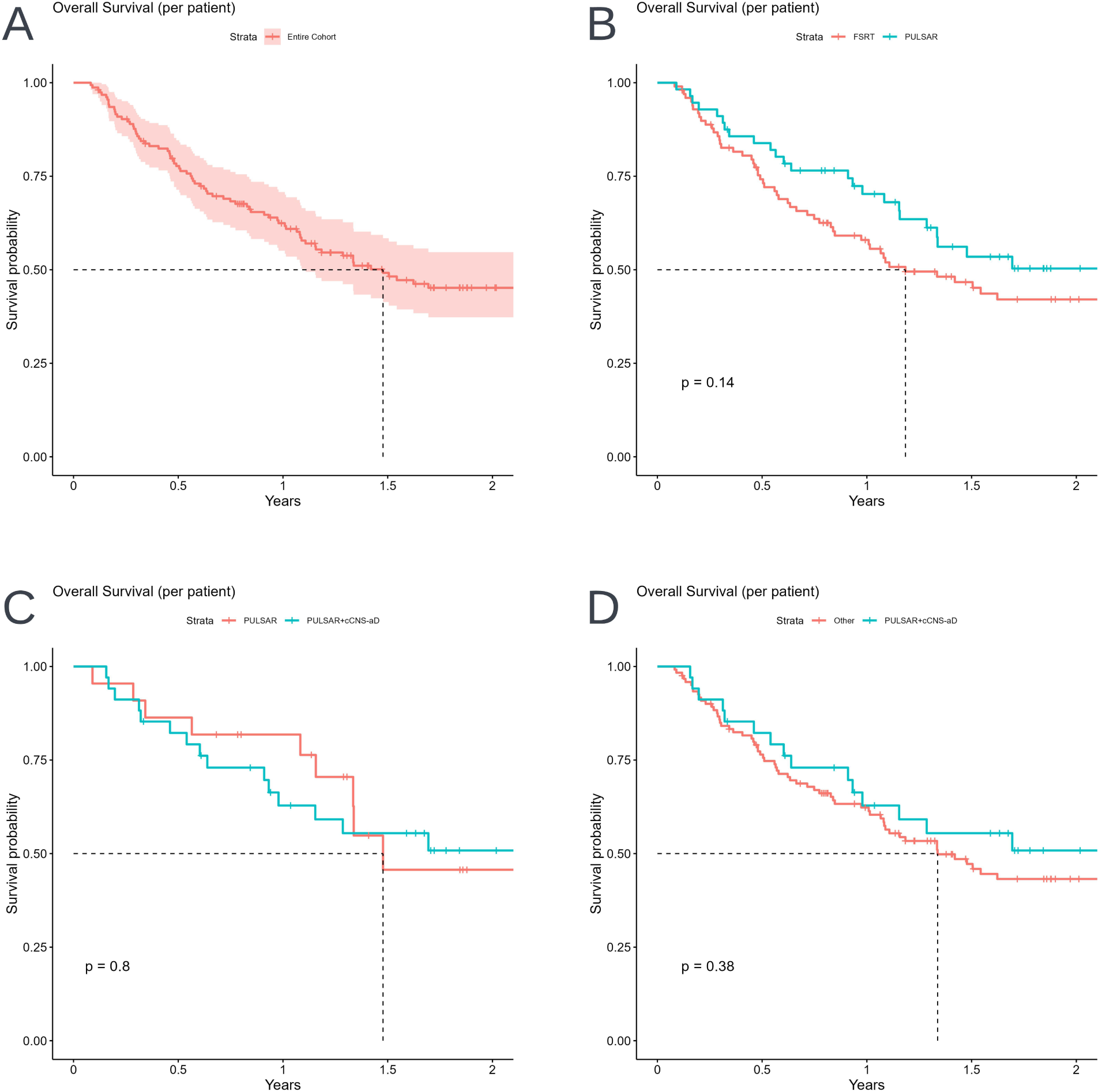
**A)** OS for the entire patient cohort. **B)** OS stratified by FSRT and PULSAR. **C)** OS of those receiving PULSAR stratified by cCNS-aD. **D)** OS stratified by treatment with PULSAR+cCNS-aD. *Abbreviations: Overall survival, OS; Fractionated stereotactic radiotherapy, FSRT; Personalized Ultrafractionated Stereotactic Adaptive Radiotherapy; PULSAR*.

